# Point-of-care C-reactive protein measurement by community health workers safely reduces antimicrobial use among children with respiratory illness: results from a stepped wedge cluster-randomized trial in rural Uganda

**DOI:** 10.1101/2024.05.22.24307733

**Authors:** Emily J. Ciccone, Di Hu, John S. Preisser, Caitlin A. Cassidy, Lydia Kabugho, Baguma Emmanuel, Georget Kibaba, Fred Mwebembezi, Jonathan J. Juliano, Edgar M. Mulogo, Ross M. Boyce

## Abstract

**Background:** Acute respiratory illness (ARI) is one of the most common reasons children receive antibiotic treatment. Measurement of C-reaction protein (CRP) to identify children at high risk for bacterial infection has been shown to reduce unnecessary antibiotic use among children with ARI in a range of clinical settings. Yet in many resource-constrained contexts, patients seek care outside the formal health sector, often from lay community health workers (CHW). The objective of this study was to determine the impact of CRP measurement on antibiotic use among children under 5 years of age presenting with febrile ARI to CHW in rural western Uganda.

**Methods and Findings:** We conducted a stepped wedge cluster randomized trial comparing a clinical algorithm that included CRP measurement by CHW to guide antibiotic treatment decisions (STAR Sick Child Job Aid [SCJA]) with the Integrated Community Care Management (iCCM) SCJA currently in use by CHW in the region. Children under 5 years presenting to CHW in 15 villages in Bugoye sub-county with fever and cough were eligible. The CHW conducted follow-up assessments 7 days after the initial visit. Our primary outcome was the proportion of children who were given or prescribed an antibiotic at the initial visit. Our secondary outcomes were (1) persistent fever on Day 7, (2) development of pre-specified danger signs, (3) unexpected visits to the CHW, (4) hospitalizations, (5) deaths, (6) lack of perceived improvement per the child’s caregiver on Day 7, and (7) clinical failure, which was a composite outcome of persistence of fever on Day 7, development of danger signs, hospitalization, or death.

The 65 participating CHW enrolled 1,280 children, 1,220 (95.3%) of whom had sufficient data to be included in this analysis; 48% (587/1,220) and 52% (633/1,220) were enrolled during control (iCCM SCJA) and intervention periods (STAR SCJA), respectively. The observed percentage of children who were given or prescribed antibiotics at the initial visit was 91.8% (539/587) in the control periods as compared to 70.8% (448/633) during the intervention periods (prevalence difference -21.0%; 95% CI: (-30.4%, - 11.7%). The odds of antibiotic prescription by the CHW were over 80% lower in the intervention as compared to the control periods (OR 0.18, 95% CI (0.07-0.45)). There was no difference in frequency of clinical failure or lack of perceived improvement by the caregiver between groups. There were no unexpected visits to the CHW or deaths in either group.

**Conclusions:** Incorporating CRP measurement into iCCM algorithms for evaluation of children with febrile ARI by CHW in rural Uganda decreased antibiotic use without negatively affecting clinical outcomes. These findings support expanded access to simple, point-of-care diagnostics to improve antibiotic stewardship in rural, resource-constrained settings where individuals with limited medical training provide a substantial proportion of care.

## Introduction

The global spread of antimicrobial resistance (AMR) represents an growing threat to child health.(1,2) A primary driver of AMR is the inappropriate use of antibiotics, which are frequently prescribed to treat pediatric febrile acute respiratory illness (ARI) in resource-constrained settings.(3–9) A large proportion of these episodes are likely caused by self-limited and/or viral infections, and therefore unlikely to need antibiotic treatment.(10,11) Yet at the same time, timely and accurate identification of the children at high risk for bacterial infection is critical to minimizing morbidity and mortality from pneumonia. Differentiating viral respiratory infections from bacterial pneumonia in resource-constrained contexts is challenging, as robust laboratory and diagnostic imaging infrastructure is not often present.

In low-and-middle-income countries (LMIC) like Uganda, lay community health workers (CHW) are assuming a greater role in the initial care of children with the goal of improving healthcare access to basic services. Many of these individuals are trained to evaluate and treat children under Integrated Community Case Management (iCCM) programs, a care strategy developed by the World Health Organization (WHO) to reduce mortality in young children related to malaria, pneumonia, and diarrhea.(12–14) Evaluations of such programs have demonstrated that CHW are able to provide a high quality of care for malaria by utilizing malaria rapid diagnostic tests (mRDTs) for diagnosis.(15–17) The quality of pneumonia management is less consistent.(15,18) According to the iCCM algorithm, children who have both cough and fast breathing (i.e., tachypnea) are diagnosed with pneumonia and treated with antibiotics. However, it is difficult to accurately measure the respiratory rate, and its assessment is influenced by contextual factors.(19,20) Furthermore, tachypnea can occur with both bacterial and viral infections. This dependence on syndromic diagnosis contributes to both (a) an overuse of antibiotics, which can drive AMR, and (b) an under-recognition of children who are at high-risk for bacterial infection.(20)

One tool for more accurately targeting antibiotic therapy is the measurement of the clinical biomarker, C-reactive protein (CRP).(21–27) CRP is released in response to inflammation, and to a greater extent in bacterial as compared to viral infection.(28) In LMIC specifically, among febrile children with ARI, clinical decision algorithms that included CRP implemented by clinicians in outpatient clinics have substantially reduced antibiotic use without increasing adverse outcomes.(29–31) However, other studies have noted a much more modest impact on antibiotic use.(26,32) Additionally, the reliance of most CRP assays on an analyzer requiring electricity and the cost of the test have limited their implementation outside of more established laboratory facilities or resource-rich contexts.

Therefore, more data on the utility of CRP to optimize the management of pediatric ARI in resource-constrained settings are needed. To date, CRP measurement to inform antibiotic decision-making in the context of iCCM care provided by lay CHW has not been evaluated, even as CHWs assume a greater role in caring for children with ARI. To address these knowledge gaps, we conducted a stepped wedge cluster-randomized trial of an adapted iCCM algorithm including point-of-care, lateral flow-based CRP measurement by CHW evaluating children with febrile ARI in rural western Uganda.

## Methods

### Study Setting

We conducted this study in the Bugoye sub-county of Kasese District in western Uganda (**Figure 1**). The region has highly variable geography; in the western villages, there are deep river valleys and steep hills of up to 2,000 meters in elevation, whereas the terrain in the eastern villages is level and at lower elevations. A quarter of the sub-county’s population of approximately 35,000 residents are children under the age of 5 years.(33)

**Figure 1.**
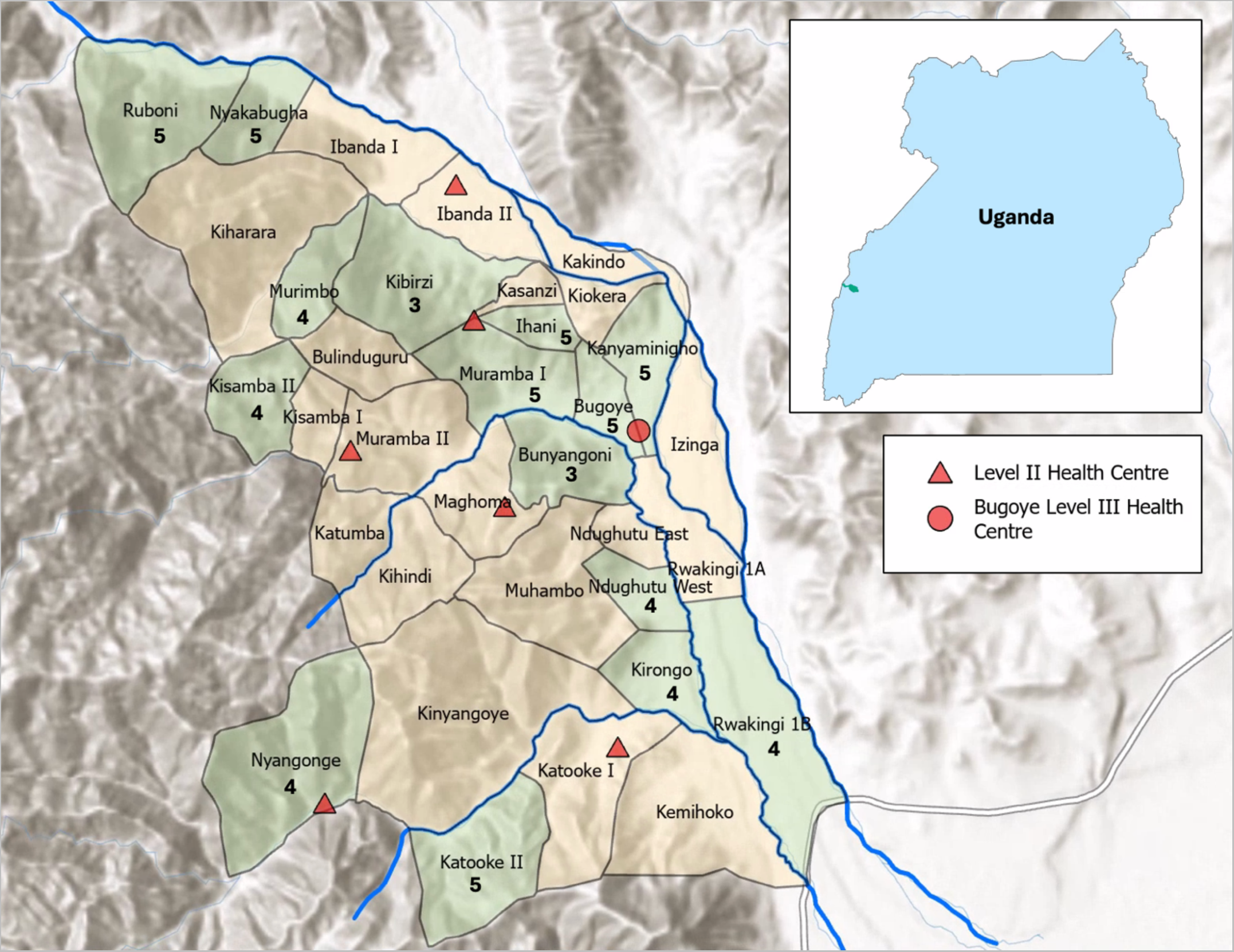
Map of Bugoye sub-county in Kasese District in western Uganda. All 34 villages within Bugoye sub-county are shown. Major rivers are shown in blue. The location of Kasese District within Uganda is shown in the inset. Villages that participated in the study are shown in green, with the number of CHWs who participated from that village. Locations of Level II and Level III health facilities indicated by red triangles and circle, respectively.

In recent years, the Government of Uganda has implemented an iCCM program consisting of teams of volunteer CHW, known as Village Health Teams (VHT). The government program began operating in Bugoye sub-county in 2013, and at the time this study was conducted in 2021-2022, external groups were also supporting additional CHW in 22 of the sub-county’s 34 villages. All CHW receive a five-day, intensive training per Ugandan Ministry of Health guidelines on receiving patients and caregivers, assessing children under age five, managing and treating diarrhea, pneumonia, and malaria, using malaria rapid diagnostic tests (mRDT), referring cases that CHW are not trained to manage, recording data, and patient follow-up. They also undergo quarterly, half-day refresher trainings. ARI, specifically defined as cough or fast breathing, is common in Bugoye sub-county – it was the second most frequent reason for a CHW visit, representing 45% of the 18,430 visits documented between April 2014 and December 2018.(34)

### Study Design

We performed a stepped-wedge, cluster randomized trial, called the Stewardship for Acute Respiratory Illness or STAR study. Villages (clusters) were switched monthly from the control to intervention in a staggered fashion. Each treatment sequence included three villages, and there were five sequences (15 villages total). The study had six observation periods of approximately one month each, with three villages randomly assigned to cross over from control to intervention at each step (**Figure 2**). We employed a cross-sectional design where different eligible children were enrolled during each month. The cross-sectional aspect meant that while individual children were in the trial for a short time (7 days of follow-up), new children were continually enrolled as they presented to the CHW.

**Figure 2.**
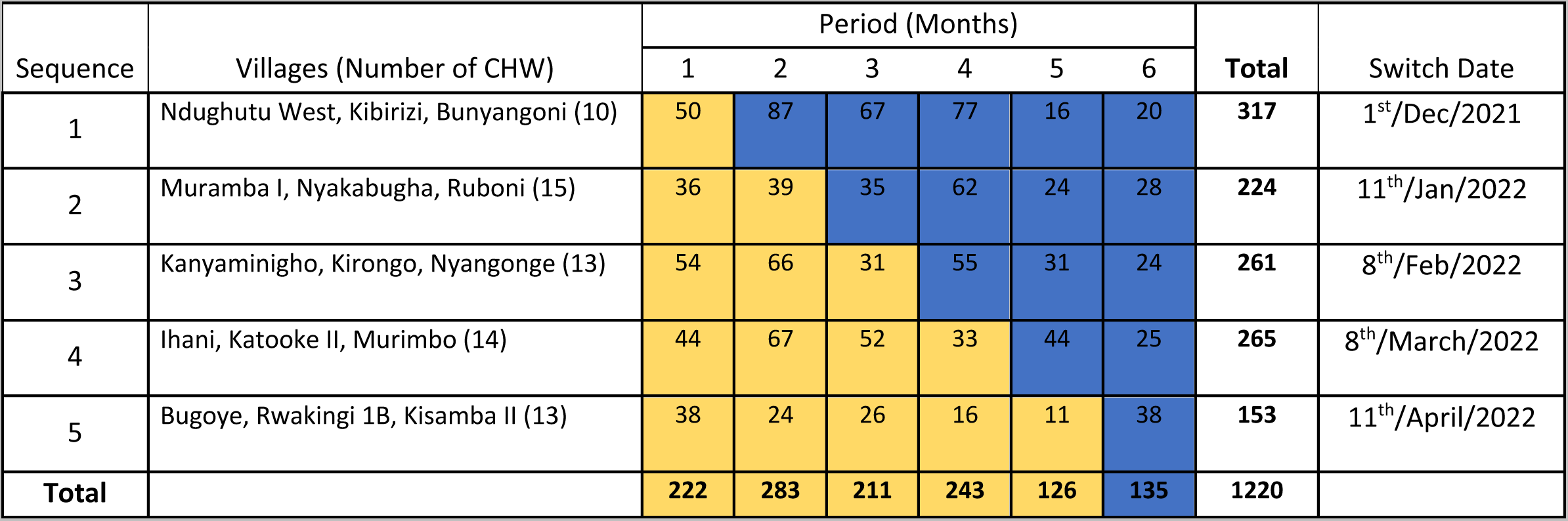
Design of the STAR stepped wedge cluster randomized trial. The study began enrollment on November 1, 2021 and ended enrollment on May 12, 2022 with each period lasting approximately 1 month. Yellow blocks represent control periods; blue blocks are intervention periods. The number shown within the blocks is the total number of participants enrolled for each sequence and period. The number of CHWs in each sequence is shown in parentheses next to the name of the villages.

### Randomization

Considering distance from Bugoye Health Centre III (red triangle in **Figure 1**), altitude, and volume of patients seen by the CHW each month, the study team selected 15 of the 22 villages with additional external support for inclusion in the trial. Each of the 15 villages selected had a robust and well-established CHW program with 3-5 CHWs per village.

To ensure balance of factors that may influence ARI epidemiology and malaria transition intensity, we performed stratified randomization by altitude, distance from the sub-county’s regional center (Bugoye), and village size, at the level of the village across treatment sequences of conditions (control v. intervention). The estimated number of eligible children seen per year by the CHW in each village was used as a proxy for village size. Emphasizing the first two factors while allowing some overlap in regard to size, we first created three strata of five villages each: (1) low altitude, proximal, “large” villages (> 115 eligible children seen per year), (2) low altitude, mid-distance, “medium size” villages (90 to 140 eligible children), and (3) high altitude, distal, “small” villages (< 120 eligible children seen). Into each of the 5 treatment sequences, we randomly selected one village from each stratum. The specifics of the villages included in each stratum are shown in Supplemental **Table S1**.

### Study Intervention

During the control periods (yellow blocks in **Figure 2**), the routine iCCM algorithm, herein referred to the iCCM Sick Child Job Aid (SCJA), was used for evaluation of enrolled children presenting with cough and fever. Per the iCCM SCJA, the CHW assessed all children for danger signs defined as severe chest in-drawing, inability to breastfeed or drink, and/or decreased level of consciousness, and if present, referred them to the nearest health facility after providing pre-referral management. If no danger signs were present, a respiratory rate was measured and, if elevated for age, antibiotic treatment for bacterial pneumonia (five days of amoxicillin) should be given (or prescribed, in the case of stockouts).

In the intervention periods (blue blocks in **Figure 2**), the CHW used a modified SCJA (herein referred to as the STAR SCJA) that included C-reactive protein (CRP) testing to guide antibiotic treatment decisions (see Supplemental **Figure S1** for full algorithm). In brief, the CHW assessed all enrolled children for danger signs and performed an mRDT and point-of-care semi-quantitative CRP test (Actim CRP, Medix Biochemica) per manufacturer instructions. If danger signs were present, the patient was referred to the nearest health facility and received pre-referral management, including antibiotics, per iCCM protocols regardless of CRP result. If no danger signs were noted, the CHW determined if antibiotic treatment for pneumonia was to be administered based on the CRP result. If CRP ≥ 40 mg/L, they dispensed five-day course of amoxicillin per local guidelines. If CRP < 40 mg/L, the CHW advised symptomatic care alone including parecetamol for fever and additional fluids to maintain hydration.

In both control and intervention periods, CHW measured mid-upper arm circumference (MUAC) for each child to assess nutritional status. They also advised all caregivers to bring the child to a health facility if, after the initial evaluation, they could not feed or drink, had a fever for more than a week, or developed chest in-drawing. They also conducted an in-person or phone follow-up assessment at 7 days with all enrolled children to assess for secondary outcomes (see below).

### Study Participants

All the CHW living in the 15 villages were approached regarding participation as study research assistants. A total of 67 CHWs underwent the initial training; two elected not to participate prior to the start of enrollment. Therefore, all children were enrolled by 65 CHW (**Figure 1**).

Children were considered eligible for the study if they were aged 2 months to 5 years, evaluated by one of the study CHW in one of the study villages, and had an acute respiratory infection defined as fever (documented (temperature > 38°C) or subjective fever in the last seven days) AND fast breathing (respiratory rate > 30 breaths per minute) OR cough. Children were excluded if a guardian was not present to provide consent. Those individuals opting not to participate underwent evaluation and treatment per iCCM guidelines (iCCM SCJA) regardless of whether the village was in a control or intervention period. Children could participate in the study more than once if the previous illness had completely resolved, and they were re-presenting for a new episode of ARI.

### Clinical Definitions

Fast breathing was a subjective assessment reported by the caregiver or the participant. Respiratory rate (RR) was determined via a manual breath count by the CHW using the timer provided by the CHW program. Tachypnea for age was defined as a RR of >50 breaths per minute (bpm) for children aged 2-11 months and >40bpm for children aged 1-5 years per local standards of care.(12,35) One CHW from each village was provided with a pulse oximeter to measure heart rate (HR) and oxygen saturation (SpO2). Tachycardia was defined as a HR of >160 beats per minute for age 2 months – 1 year, >150 for ages 1-3 years, and >140 for ages 3-5 years per the WHO.(36) Hypoxia was defined as an SpO2 of less than 90%. Severe acute malnutrition and moderate acute malnutrition were defined as a MUAC of <115 mm and ≥115 mm to <125 mm, respectively, per WHO standards.(37)

### Outcomes

The primary child-level outcome was whether an antibiotic was given or prescribed by the CHW during the initial assessment of the child. In addition, we performed a post-hoc subgroup analysis of our primary outcome by mRDT result status. Finally, we conducted an ancillary analysis of whether an antibiotic was given or prescribed either at the initial (baseline) visit or during the follow-up period (which was a pre-specified secondary outcome).

There were seven pre-specified secondary outcomes: (1) persistent fever on Day 7, (2) development of danger signs, (3) unexpected visits to the CHW during the 7-day follow-up period, (4) hospitalizations, (5) deaths, (6) lack of perceived improvement per the child’s caregiver on Day 7, and (7) clinical failure, which was a composite outcome of persistence of fever at the follow-up assessment, development of danger signs, hospitalization, or death. Posthoc secondary outcomes included (1) an alternate clinical failure composite outcome of lack of perceived improvement by caregiver on Day 7, development of danger signs, and/or hospitalization, (2) a composite outcome of need for further outpatient evaluation during the study follow-up period (visit to a traditional healer, drug shop, or clinic for the same problem), (3) the persistence of symptoms other than fever at Day 7, and (4) presence of tachypnea at Day 7.

### Statistical Methods

Descriptive statistics were used to calculate the proportion of children who were given or prescribed antibiotics by treatment condition and by sequence-period. The analysis of the primary outcome employed a marginal logistic regression model estimated within a generalized estimating equations (GEE) framework to describe how the probability of antibiotic use changed across the subsets of the population defined by the intervention and control cluster-periods (see Supplemental File for details). The analysis performed with the SAS macro GEEMAEE (39) treated villages as clusters, fitted a logistic model that included period as a categorical variable and an indicator variable for intervention; specified a within-village, nested exchangeable correlation structure having one within-period and one between-period correlation parameter to account for correlation decay over time; and made two kinds of small-sample bias-corrections considering that 15 clusters (villages) is too small a number to otherwise reliably employ standard GEE. Thus, the analysis used bias-corrected variance estimators for model parameter estimates, and bias corrections to the estimated within-cluster correlations via a procedure known as matrix-adjusted estimating equations (MAEE).(38,39) From the logistic model, the odds ratio for the intervention effect and its 95% confidence interval were computed (Supplemental File). Similar models for prevalence differences are also reported, as are results of the subgroup and ancillary analyses for antibiotic use. There were only three participants enrolled during an intervention period who were managed as in the control condition (using the iCCM SCJA). As this number was so small, these individuals were included as-treated. Analyses were performed using SAS 9.4 and R 4.3.1.

For secondary outcomes, which were uncommon events in that they occurred in less than 10% of study participants, we used a stratified logistic regression model with linear period effects, the intervention indicator variable and village as fixed effects. We used exact conditional inference with the EXACT statement in SAS Proc Logistic with village managed as a stratification factor. An exception was tachypnea; it was analyzed with GEE/MAEE like the primary outcome since it was not an uncommon outcome.

### Power calculation and sample size

Prior to initiation of the trial, a statistical power formula for stepped wedge designs based on GEE analysis was applied for the dichotomous primary outcome, assuming 15 clusters (villages) transitioning from control to intervention condition in five waves of 3 clusters each and 6 total periods of follow-up, as in **Figure 2**, except that an equal number of children were assumed in each sequence-period (38) (40)Assuming twelve children recruited per village-month (total sample size of 1080), an antibiotic use rate of 80% under the control condition and 60% under the intervention condition, the study had 87% power corresponding to an odds ratio of 0.375 with two-sided α=0.05 GEE Wald tests assuming a zero-slope temporal trend, a within-period correlation of 0.10 and a between-period correlation of 0.05. Post-hoc power, based on the observed unequal cluster-period sizes in **Figure S2** and actual enrollment of 1220 was 97% (see Supplemental File).

### Research Ethics Review and Trial Registration

The study was reviewed and approved by the University of North Carolina Institutional Review Board (#18-2803), Mbarara University of Science and Technology Research Ethics Committee (14/03-19), and Uganda National Council on Science and Technology (HS 2631). Study enrollment took place between November 1, 2021 and May 12, 2022. Due to an administrative error, trial registration occurred after recruitment began (ClinicalTrials.gov NCT05294510; March 23, 2022). For detailed timeline of ethics review, trial registration, and participant approval, please see Supplemental Regulatory Materials.

## Results

### Study Population

A total of 1,280 children were enrolled between November 2021 and May 2022 (**Figure 3**). Sixty individuals were excluded from this analysis because of missing data collection forms. Slightly more participants were enrolled during intervention periods (n=633, 52%) as compared to the control periods (n=587, 48%).

**Figure 3.**
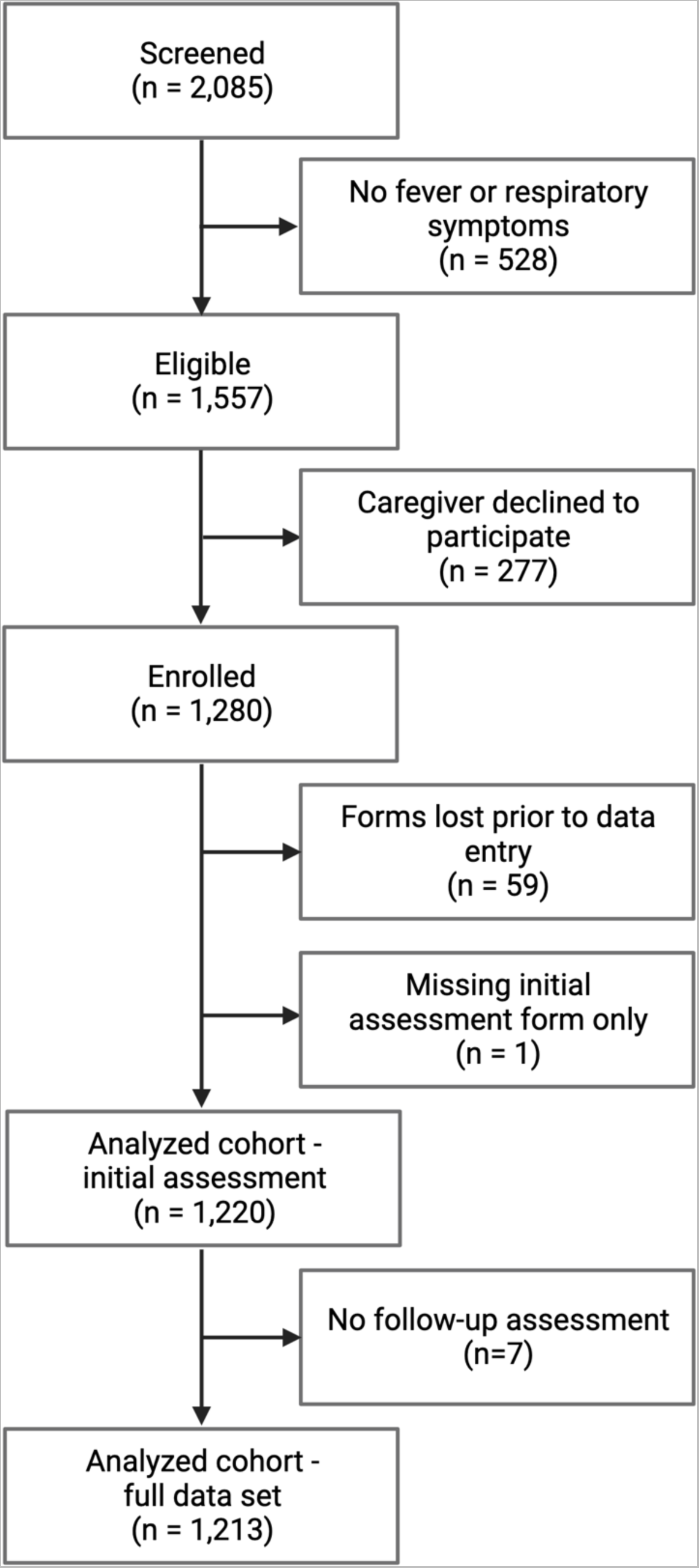
Participant Flow Diagram.

The number of children enrolled in each period is shown by sequence (**Figure 2**) and by village (Supplemental **Figure S2**). The number of children enrolled per CHW over the course of the study ranged from 3 to 49 with a median of 16.5 (Supplemental **Figure S3**). The CHW in villages from the first treatment sequence enrolled the highest average number of participants per person (mean (standard deviation (sd)) = 31.7 (9.2)) whereas the CHWs in villages from the last (fifth) sequence had the lowest (mean (sd) = 11.8 (7.3)). Fewer participants were enrolled in the last two periods than the first four periods (**Figures 2 and S2**).

### Baseline Characteristics

Demographic characteristics of the study participants are shown in **Table 1**. Of note, age, sex, and family size were similar for children enrolled during control and intervention periods. A slightly higher proportion of children lived in permanent houses, and a slightly lower proportion lived in semi-permanent or temporary structures, in the control as compared to the intervention group. The distribution of the village size of the enrollees was similar in the control and intervention groups, with a slightly higher proportion of children living in small villages in the intervention group.

**Table 1.** Demographics of the study population. All values show are n(%) unless otherwise indicated.

|  | **Control**<br>**(N=587)** | **Intervention**<br>**(N=633)** | **Total**<br>**(N=1220)** |
| --- | --- | --- | --- |
| **Age in months** |  |  |  |
| Median (IQR) | 24.0 (12.0, 36.0) | 24.0 (15.0, 36.0) | 24.0 (13.0, 36.0) |
| **Sex** |  |  |  |
| Female | 310 (52.8%) | 329 (52.0%) | 639 (52.4%) |
| **Occupation of guardian** |  |  |  |
| Peasant | 563 (95.9%) | 588 (93.0%) | 1151 (94.4%) |
| Teacher | 10 (1.7%) | 15 (2.4%) | 25 (2.1%) |
| Businessperson | 4 (0.7%) | 9 (1.4%) | 13 (1.1%) |
| Boda boda driver | 6 (1.0%) | 4 (0.6%) | 10 (0.8%) |
| Tailor | 1 (0.2%) | 9 (1.4%) | 10 (0.8%) |
| Student | 2 (0.3%) | 1 (0.2%) | 3 (0.2%) |
| Trader | 0 (0.0%) | 3 (0.5%) | 3 (0.2%) |
| Civil Servant | 0 (0.0%) | 1 (0.2%) | 1 (0.1%) |
| Engineer | 1 (0.2%) | 0 (0.0%) | 1 (0.1%) |
| LC1 chairman | 0 (0.0%) | 1 (0.2%) | 1 (0.1%) |
| Community Health Worker | 0 (0.0%) | 1 (0.2%) | 1 (0.1%) |
| **Family Size** |  |  |  |
| Median (IQR) | 6.0 (4.0, 8.0) | 6.0 (5.0, 8.0) | 6.0 (5.0, 8.0) |
| **Home Construction** |  |  |  |
| Permanent (bricks with iron sheets) | 278 (47.4%) | 242 (38.2%) | 520 (42.7%) |
| Semi-permanent (mud with iron sheets) | 282 (48.1%) | 352 (55.6%) | 634 (52.0%) |
| Temporary (mud with grass thatched roof) | 26 (4.4%) | 39 (6.2%) | 65 (5.3%) |
| Missing | 1 | 0 | 1 |
| **Village Stratum** |  |  |  |
| Large | 245 (41.7%) | 234 (37.0%) | 479 (39.3%) |
| Medium | 177 (30.2%) | 185 (29.2%) | 362 (29.7%) |
| Small | 165 (28.1%) | 214 (33.8%) | 379 (31.1%) |

The clinical characteristics of the participants are described in **Table 2**. Fewer participants and their caregivers in the intervention arm reported fast breathing as a symptom, whereas more had a fever documented by the clinician during the initial assessment. The proportion of children with tachypnea for age as measured by the CHW was similar in both groups, as was the frequency with which cough was reported. Among the subset of participants for whom the study CHW measured HR and SpO2, a slightly higher proportion were tachycardic for age in the intervention arm. The distribution of nutritional status was similar in both groups.

**Table 2.**
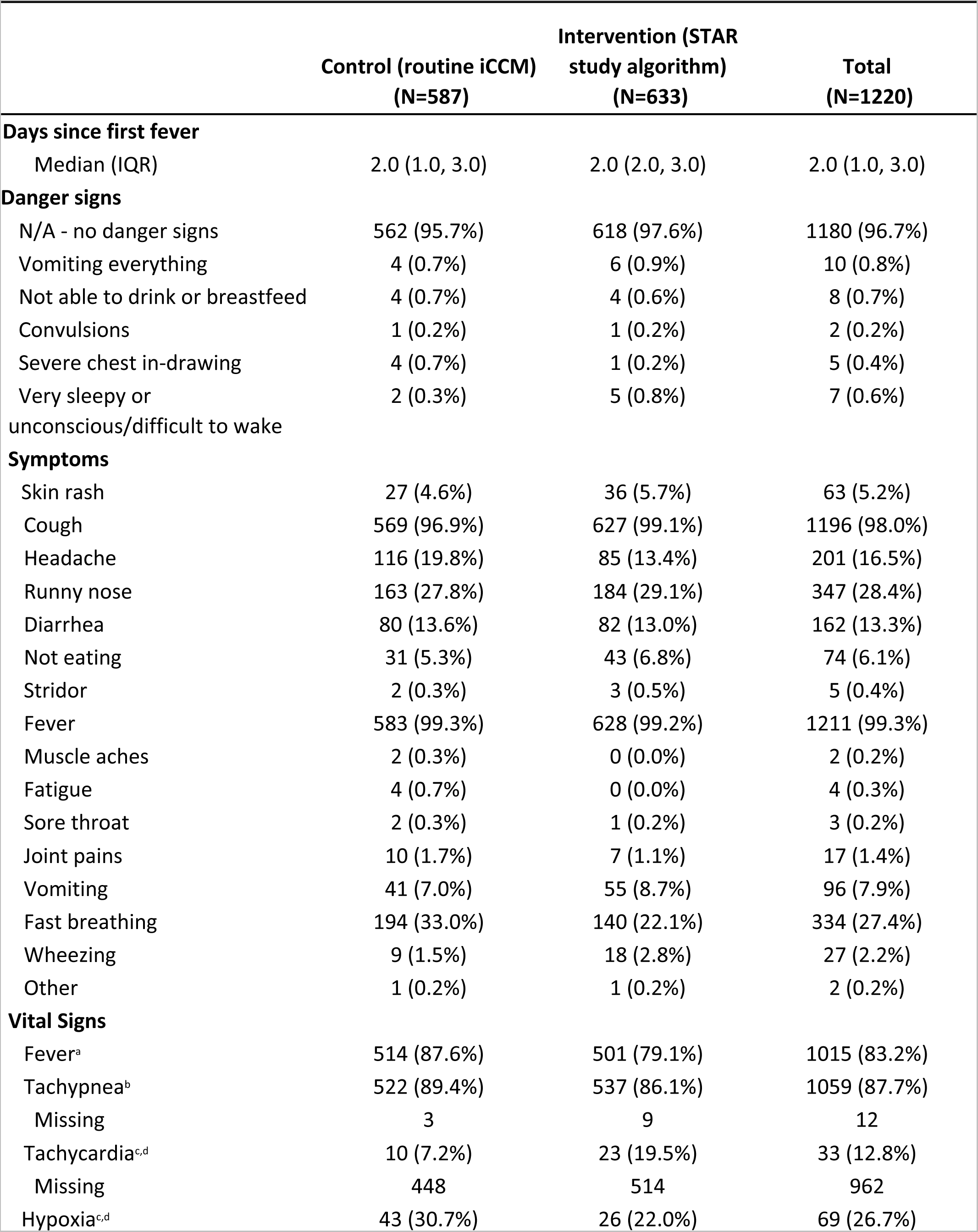
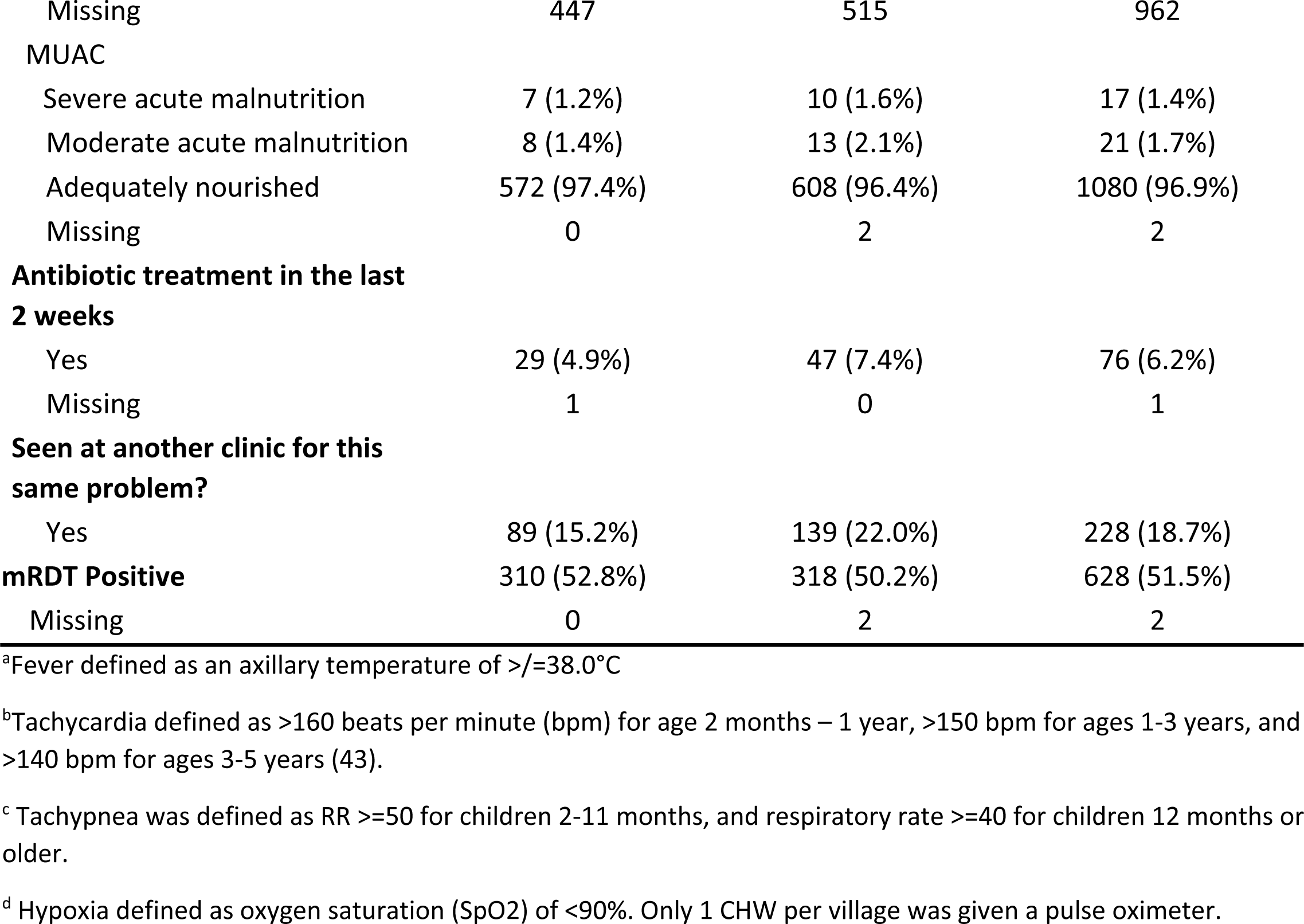
Clinical presentation of the study participants. All values show are n(%) unless otherwise indicated.

|  | Control (routine iCCM)<br>(N=587) | Intervention (STAR<br>study algorithm)<br>(N=633) | Total<br>(N=1220) |
| --- | --- | --- | --- |
| **Days since first fever** |  |  |  |
| Median (IQR) | 2.0 (1.0, 3.0) | 2.0 (2.0, 3.0) | 2.0 (1.0, 3.0) |
| **Danger signs** |  |  |  |
| N/A - no danger signs | 562 (95.7%) | 618 (97.6%) | 1180 (96.7%) |
| Vomiting everything | 4 (0.7%) | 6 (0.9%) | 10 (0.8%) |
| Not able to drink or breastfeed | 4 (0.7%) | 4 (0.6%) | 8 (0.7%) |
| Convulsions | 1 (0.2%) | 1 (0.2%) | 2 (0.2%) |
| Severe chest in-drawing | 4 (0.7%) | 1 (0.2%) | 5 (0.4%) |
| Very sleepy or<br>unconscious/difficult to wake | 2 (0.3%) | 5 (0.8%) | 7 (0.6%) |
| **Symptoms** |  |  |  |
| Skin rash | 27 (4.6%) | 36 (5.7%) | 63 (5.2%) |
| Cough | 569 (96.9%) | 627 (99.1%) | 1196 (98.0%) |
| Headache | 116 (19.8%) | 85 (13.4%) | 201 (16.5%) |
| Runny nose | 163 (27.8%) | 184 (29.1%) | 347 (28.4%) |
| Diarrhea | 80 (13.6%) | 82 (13.0%) | 162 (13.3%) |
| Not eating | 31 (5.3%) | 43 (6.8%) | 74 (6.1%) |
| Stridor | 2 (0.3%) | 3 (0.5%) | 5 (0.4%) |
| Fever | 583 (99.3%) | 628 (99.2%) | 1211 (99.3%) |
| Muscle aches | 2 (0.3%) | 0 (0.0%) | 2 (0.2%) |
| Fatigue | 4 (0.7%) | 0 (0.0%) | 4 (0.3%) |
| Sore throat | 2 (0.3%) | 1 (0.2%) | 3 (0.2%) |
| Joint pains | 10 (1.7%) | 7 (1.1%) | 17 (1.4%) |
| Vomiting | 41 (7.0%) | 55 (8.7%) | 96 (7.9%) |
| Fast breathing | 194 (33.0%) | 140 (22.1%) | 334 (27.4%) |
| Wheezing | 9 (1.5%) | 18 (2.8%) | 27 (2.2%) |
| Other | 1 (0.2%) | 1 (0.2%) | 2 (0.2%) |
| **Vital Signs** |  |  |  |
| Fever<sup>a</sup> | 514 (87.6%) | 501 (79.1%) | 1015 (83.2%) |
| Tachypnea<sup>b</sup> | 522 (89.4%) | 537 (86.1%) | 1059 (87.7%) |
| Missing | 3 | 9 | 12 |
| Tachycardia<sup>c,d</sup> | 10 (7.2%) | 23 (19.5%) | 33 (12.8%) |
| Missing | 448 | 514 | 962 |
| Hypoxia<sup>c,d</sup> | 43 (30.7%) | 26 (22.0%) | 69 (26.7%) |
| Missing | 447 | 515 | 962 |
| **MUAC** |  |  |  |
| Severe acute malnutrition | 7 (1.2%) | 10 (1.6%) | 17 (1.4%) |
| Moderate acute malnutrition | 8 (1.4%) | 13 (2.1%) | 21 (1.7%) |
| Adequately nourished | 572 (97.4%) | 608 (96.4%) | 1080 (96.9%) |
| Missing | 0 | 2 | 2 |
| **Antibiotic treatment in the last 2 weeks** |  |  |  |
| Yes | 29 (4.9%) | 47 (7.4%) | 76 (6.2%) |
| Missing | 1 | 0 | 1 |
| **Seen at another clinic for this same problem?** |  |  |  |
| Yes | 89 (15.2%) | 139 (22.0%) | 228 (18.7%) |
| **mRDT Positive** | 310 (52.8%) | 318 (50.2%) | 628 (51.5%) |
| Missing | 0 | 2 | 2 |

<sup>a</sup>Fever defined as an axillary temperature of $\geq 38.0^{\circ}\text{C}$
<sup>b</sup>Tachycardia defined as $>160$ beats per minute (bpm) for age 2 months – 1 year, $>150$ bpm for ages 1-3 years, and $>140$ bpm for ages 3-5 years (43).
<sup>c</sup> Tachypnea was defined as RR $\geq 50$ for children 2-11 months, and respiratory rate $\geq 40$ for children 12 months or older.
<sup>d</sup> Hypoxia defined as oxygen saturation (SpO<sub>2</sub>) of $<90\%$ . Only 1 CHW per village was given a pulse oximeter.

### Antibiotic Use

Our primary outcome of interest was antibiotic treatment (amoxicillin) given or prescribed by the CHW at the initial visit (**Table 3**). In the control periods, CHWs gave antibiotics to 91.8% (539/587) of children as compared to 70.8% (448/633) during the intervention periods (prevalence difference -21.0% (95% CI: -30.4%, -11.7%)). The observed proportions of antibiotic use varied slightly over time but with no apparent temporal trend (Supplemental **Figure S4**). After adjusting for month of visit, the odds of antibiotic prescription by the CHW were over 80% lower in the intervention as compared to the control periods (OR 0.18, 95% CI (0.07-0.45)). We observed a similar trend in both mRDT-positive and mRDT-negative children (**Table 3**). Full results of the logistic regression model for antibiotic use are given in Supplemental **Table S2**.

**Table 3.** Comparison of antibiotic use among children under the control and intervention treatment conditions during the STAR stepped wedge cluster randomized trial (as treated analysis). Children seen during control periods were evaluated and managed using the iCCM SCJA; children seen during the intervention condition were evaluated and managed using the STAR SCJA. Three children seen during an intervention period were evaluated using the iCCM SCJA; they were included in the control condition.

|  |  | Observed Proportions |  | Intervention Effect<sup>1</sup> (95% CI) |  |
| --- | --- | --- | --- | --- | --- |
|  |  | Control | Intervention | Prevalence Difference | Odds Ratio |
| Antibiotics at initial visit | All children | 539/587<br>(91.8%) | 448/633<br>(70.8%) | -21.0%<br>(-30.4%, -11.7%) | 0.18<br>(0.07, 0.45) |
|  | mRDT+ | 285/310<br>(91.9%) | 228/318<br>(71.7%) | -20.2%<br>(-28.5%, -12.0%) | 0.18<br>(0.06, 0.56) |
|  | mRDT - | 254/277<br>(91.7%) | 218/313<br>(69.6%) | -22.1%<br>(-32.9%, -11.2%) | 0.18<br>(0.06, 0.55) |
| Antibiotics at any time |  | 540/584<br>(92.5%) | 458/620<br>(73.9%) | -18.6%<br>(-26.9%, -10.3%) | 0.18<br>(0.06, 0.48) |

<sup>1</sup>Results based on GEE assuming marginal Bernoulli distributions for any antibiotic use and bias-corrected standard errors to adjust for the moderately small number of clusters (villages). The prevalence difference is based on an identity link generalized linear model with intercept and an intervention indicator variable with working independence correlation. The odds ratio is based on a logit link (i.e., logistic regression) model with intercept, categorical periods effects, and an intervention indicator variable with a nested exchangeable working correlation structure, and correlation parameters (see supplement including Table S2) estimated with MAEE.

Among those who did not receive antibiotics at the initial visit, there were numerically more participants in the intervention group who acquired antibiotics between the initial and follow-up visits, but the absolute number of additional antibiotic prescriptions was low in both groups ((10/172, 5.8% v. 1/45, 2.2%; OR 2.32 (95% CI 0.19-130.38)). As such, analysis results of the secondary outcome of antibiotics given or prescribed at any point (initial visit or during follow-up) among children for whom data were available at both time points, were like that of the primary outcome.

### Clinical Failure and Adverse Events

Secondary outcomes assessing the safety of the STAR algorithm are detailed in **Figure 4** and Supplemental **Table S3**. Overall, we did not find evidence to suggest any relationship between the intervention and secondary outcomes as all 95% confidence intervals for odds ratios included 1.0. Specifically, clinical failure was uncommon and occurred at a slightly lower frequency in the intervention group (control 3.9% (23/585) v. intervention 1.8% (11/630); OR 0.41, 95% CI 0.09-1.83). There was no difference in the risk of hospitalization or persistence of fever, both of which were rare events. There were numerically fewer participants with any danger sign at the follow-up visit in the intervention arm, but this outcome was overall rare in both groups (1.7% (10/585) v. 0.5% (3/630); OR 0.32, 95% CI 0.02-5.12). No unexpected visits to the CHW between initial and follow-up visits occurred, and there were no deaths during study follow-up. Few caregivers perceived that their child had not improved at the Day 7 follow-up assessment (control: 12/584 (2.1%) v. intervention: 22/627 (3.5%) OR 1.49, 95% CI 0.37-6.52).

**Figure 4.**
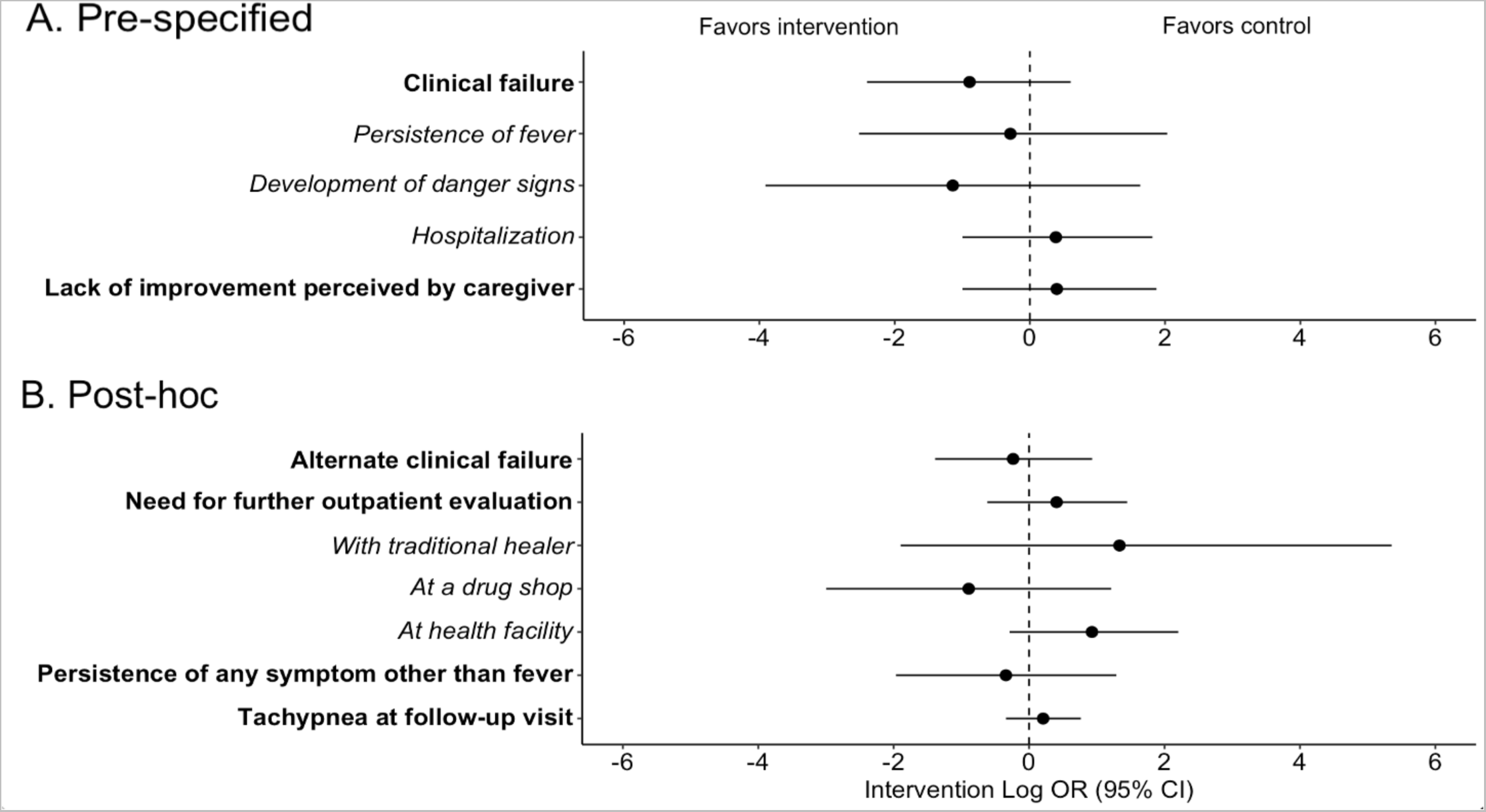
Forest plot of secondary outcomes. A. Pre-specified secondary outcomes. B. Post-hoc secondary outcomes. Odds ratios presented on the log scale with point estimates shown by circles and 95% confidence intervals shown by error bars. Dashed line indicates null value. Individual components of composite outcomes indicated with italic text below the corresponding composite outcome.

To further assess the impact and safety of the intervention, we conducted several post-hoc analyses of clinical outcomes and care-seeking (**Figure 4**, Supplemental **Table S3**). Among those who had a respiratory rate measured at the follow-up visit, the proportion who were tachypneic for age was similar between control and intervention conditions (70/414 (16.9%) v. 78/460, (17.0%); OR 1.23, 95% CI 0.71-2.15). Persistence of symptoms other than fever was rare and occurred at a similar frequency in both conditions (11/585, 1.9% v. 14/630, 2.2%; OR 0.71 (0.14-3.63). Finally, there was no difference in further care-seeking from another outpatient provider (traditional healer, drug shop, or outpatient clinic) for the same illness (control: 32/585, 5.5% v. intervention: 33/630, 5.2%; OR 1.50, 95% CI 0.54-4.26).

### CRP Test Results

The distribution of CRP testing results in the intervention group is shown in **Figure 5**. The majority of participants had CRP levels >40 mg/L (444/631, 70.3%), with the most common test result being 40-80mg/L (371/631, 58.8%). Only 11.6 % of participants had CRP levels of >80 mg/L.

**Figure 5.**
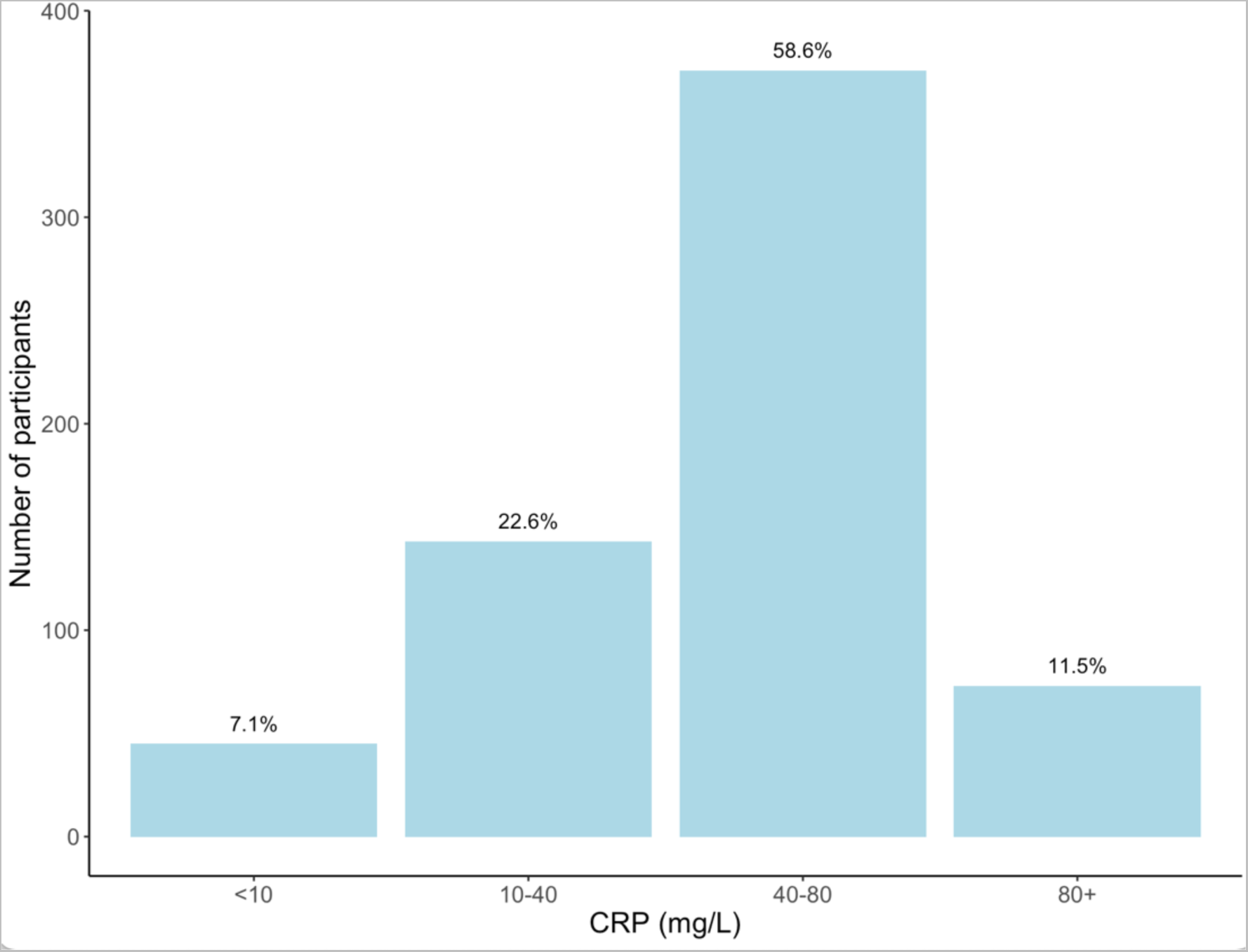
Distribution of C-reactive protein measurements from children enrolled during the intervention periods.

### Adherence to Algorithm

Antibiotic prescriptions by CRP test result are shown in **Table 4**. Of those with CRP < 40mg/L, 1.6% (3/631) received antibiotics despite the STAR SCJA advising against it. A CHW noted that for one of these participants, the caregiver insisted on an antibiotic prescription.

**Table 4.** Algorithm adherence for intervention (CRP, top) and control (tachypnea and cough, bottom) groups.^1^.

|  | **Antibiotic Use** |  |  |
| --- | --- | --- | --- |
| **CRP (mg/L)** | **No** | **Yes** | **Total** |
| **<40**<br>**n (%)** | 185 (98.4%) | 3 (1.6%) | 188 |
| **40+**<br>**n (%)** | 0 (0%) | 444 (100%) | 444 |
| **Total** | 185 | 447 | 632 |
|  | **Antibiotic Use** |  |  |
| **Tachypnea<sup>2</sup>**<br>**and cough** | **No**<br>**n (%)** | **Yes**<br>**n (%)** | **Total** |
| **No**<br>**n (%)** | 38 (49.4%) | 39 (50.6%) | 77 |
| **Yes**<br>**n (%)** | 10 (2.0%) | 497 (98.8%) | 507 |
| **Total** | 48 | 536 | 584 |

<sup>1</sup>CRP measurements were missing for one participant and respiratory rate (RR) was missing for three children.
<sup>2</sup>Tachypnea was defined as RR ≥ 50 for children 2-11 months, and respiratory rate ≥ 40 for children 12 months or older as per the iCCM SCJA.

Per the iCCM SCJA, elevated respiratory rate for age determines which children with cough should be given antibiotics. Among participants in the control condition without tachypnea and cough, 50.6% (39/77) received antibiotics. There were also 10 children (2%) who were tachypneic and had cough who did not receive antibiotics, six of whom were mRDT-positive.

### Associated Costs

Purchasing the CRP tests directly from the manufacturer, the cost per test was 1.53 EUR (approximately 1.70 USD). Assuming a 20% decrease in antibiotic use as observed in this study, the number needed to test to avoid one course of antibiotics would be 5 with an associated cost of $8.50. A course of amoxicillin treatment for a child aged 1-5, if purchased from a local drug shop, ranges from 2,000-5,000 UGX (approximately 0.50-1.30 USD) depending on country of manufacture.

## Discussion

This article reports results from a stepped wedge cluster randomized trial of an adaptation of the iCCM algorithm, the STAR SCJA, that employs point-of-care CRP measurement by CHW to guide antibiotic treatment decisions for children presenting with febrile ARI in rural western Uganda. As compared to the standard iCCM algorithm, the use of the STAR SCJA was associated with a significant decrease in antibiotic use without a concomitant increase in adverse outcomes. Importantly, this impact was achieved by non-medically trained, volunteer health workers, who are often the first providers from whom children seek care in this region. To our knowledge, this is the first study to demonstrate that CHW can implement and interpret point-of-care CRP testing as part of a broader algorithm.

The magnitude of the impact of CRP measurement on antibiotic use was similar to what has been observed in primary care clinics in Vietnam,(21,26) but less than what has been seen in neighboring Tanzania.(30) The reasons for this are likely several fold. We intentionally chose a lower CRP cut-off for antibiotic prescription (40mg/L) than what was used in the Tanzanian study (80mg/L) to favor sensitivity for bacterial infection over specificity as the CHW are not formally medically trained. This choice of cut-off was also supported by work demonstrating a CRP cut-off of 44.1mg/L as having a negative predictive value of 92% for radiologic end-point pneumonia,(22) and a trial conducted in Myanmar and Thailand in which a cut-off of 40mg/L for antibiotic prescription in febrile illness was safe.(32) In our study, only 12% of participants had CRP levels of >80 mg/L. Therefore, assuming complete adherence by providers to the algorithm, had that higher cut-off been used, we would have observed similar decreases in antibiotic use as the Tanzanian study. In addition, in the Tanzanian study, the CRP test was included as part of a package of multiple rapid diagnostics in the context of an electronic clinical decision support algorithm for all pediatric febrile illness that, for evaluation of children with respiratory illness, also incorporated respiratory rate thresholds that accounted for age and temperature. The incorporation of multiple stewardship interventions may have led to additional decreases in antibiotic use. Finally, a higher proportion of children were mRDT-positive in this study than anticipated based on our previous research in the region and as compared to other similar studies.(30) As malaria infection itself is associated with increased CRP levels,(41) this could have contributed to the high proportion of children with CRP levels 40-80mg/L (∼60%) who received antibiotics per protocol.

Interestingly, a study in Myanmar and Thailand observed a smaller absolute difference in antibiotic prescribing (risk difference of 5%) using the same CRP cut-off as our study. This disparity may be due to differences in study population (it included both children and adults presenting with any febrile illness to primary care clinics) and lower baseline frequency of antibiotic prescribing (39% in the control group). In addition, in comparison to this and other pragmatic evaluations of CRP use, we observed very high uptake of CRP use and adherence to the study algorithm in our study.(26,29,31) Our research group has been conducting studies with the CHW in this region for 10 years, and overall, community perception of health-related interventions is positive. The local CHW have previously demonstrated high adherence to routine iCCM algorithms,(18,34) and anecdotally, the CHWs reported confidence in the CRP test and appreciation of having an additional tool to inform their treatment decisions. We conducted a qualitative sub-study of CHW who participated in the study to examine this further, the results of which will be published separately.

Importantly, the decrease in antibiotic use that we noted in the intervention condition was not associated with an increase in adverse clinical outcomes. Although the study was not designed to assess non-inferiority of the STAR SCJA for clinical failure (a rare outcome), our results are consistent with numerous other studies that have demonstrated the safety of CRP testing for antibiotic stewardship.(21,26,29–31) Future studies of CRP use by CHW should be powered to detect differences in clinical outcomes and more rigorously assess cost-effectiveness by incorporating both costs associated with avoidance of unnecessary antibiotic use, potential decrease in adverse clinical outcomes, and impact on antimicrobial resistance.

Our study had numerous strengths. By adding the CRP test to routine iCCM care provided by non-professional CHWs, we extend the previous research in health facilities to a novel context – one which represents the first point of healthcare contact for many caregivers with ill children in rural areas of sub-Saharan Africa. Furthermore, the cluster-randomized design likely minimized risk of between group contamination and the Hawthorne effect as was a concern in previous work.(32) Adherence to the STAR SCJA was high, and there was minimal loss to follow-up. Finally, our study used novel statistical methods for population-averaged models in an integrated approach to study design (and power) and analysis of dichotomous outcomes (with generalized estimating equations) in a stepped wedge design with a moderately small number of clusters (15 villages).

The study, however, also had limitations. First, as described in the Methods section and Supplemental Materials, the trial was not prospectively registered, which introduces the potential for investigator bias. However, the risk of this bias is thought to be minimal as the study protocol and analysis plan were pre-specified and received ethical approval prior to start of enrollment (see Supplemental Materials), the trial was registered prior to the completion of enrollment, and information regarding all enrolled participants is included in this paper. In addition, there were some data collection forms that were lost prior to data entry. Despite this, our sample size exceeded our enrollment estimates, so we still had sufficient power to detect a difference between groups (see Supplemental File); therefore, these missing data very likely would not have had a significant impact on the study findings. We also did not systematically evaluate CHW performance of the CRP test in comparison with an expert medical laboratory technician. Finally, further studies are needed to assess feasibility and acceptability of the STAR SCJA and CRP test when implemented by community health worker programs elsewhere in Uganda and in other countries to ensure our findings are broadly generalizable.

In conclusion, the addition of point-of-care CRP measurement to routine iCCM care by CHW is feasible and has the potential to reduce unnecessary antibiotic use outside of the formal health sector. These findings support expanded, more equitable, access to simple diagnostics to improve antibiotic stewardship in rural, resource-constrained settings where individuals with limited medical training are often the first providers from whom ill children seek care.

## Data Availability

Deidentified individual data that support the results will be shared following publication provided the investigator who proposes to use the data has approval from an Institutional Review Board (IRB), Independent Ethics Committee (IEC), or Research Ethics Board (REB), as applicable, and executes a data use/sharing agreement with UNC. Researchers may apply for data access by contacting the corresponding author or the UNC IRB at.

## Acknowledgements

We sincerely thank the Village Health Teams for their enthusiastic implementation of this study. We are also greatly appreciative of all the children and caregivers who participated in the study and the local community leaders for their support of this work.

